# Artificial intelligence-driven real-time 3D surface quantification of Barrett’s oesophagus for risk stratification and therapeutic response monitoring

**DOI:** 10.1101/2020.10.04.20206482

**Authors:** Sharib Ali, Adam Bailey, James E. East, Simon J. Leedham, Maryam Haghighat, TGU Investigators, Xin Lu, Jens Rittscher, Barbara Braden

## Abstract

**BACKGROUND & AIMS:** Barrett’s epithelium measurement using widely accepted Prague C&M criteria is highly operator dependent. By reconstructing the surface of the Barrett’s area in 3D from endoscopy video, we propose a novel methodology for measuring the C&M score automatically. This 3D reconstruction provides an extended field of view and also allows to precisely quantify the Barrett’s area including islands. We aim to assess the accuracy of the extracted measurements from phantom and demonstrate their clinical usability.

**METHODS:** Advanced deep learning techniques are utilised to design estimators for depth and camera pose required to map standard endoscopy video to a 3D surface model. By segmenting the Barrett’s area and locating the position of the gastro-oesophageal junction (GEJ) we measure C&M scores and the Barrett’s oesophagus areas (BOA). Experiments using a purpose-built 3D printed oesophagus phantom and high-definition video from 98 patients scored by an expert endoscopist are used for validation.

**RESULTS:** Endoscopic phantom video data demonstrated a 95 % accuracy with a marginal ± 1.8 mm average deviation for C&M and island measurements, while for BOA we achieved nearly 93 % accuracy with only ± 1.1 cm^2^ average deviation compared to the ground-truth measurements. On patient data, the C&M measurements provided by our system concord with the reference provided by expert upper GI endoscopists.

**CONCLUSIONS:** The proposed methodology is suitable for extracting Prague C&M scores automatically with a high degree of accuracy. Providing an accurate measurement of the entire Barrett’s area provides new opportunities for risk stratification and the assessment of therapy response.

## Introduction

Barrett’s oesophagus is a precancerous condition associated with an annual progression rate to oesophageal adenocarcinoma (EAC) at 0.12–0.13% per year.^1-2^ In patients with this condition, the oesophageal squamous mucosa is replaced by columnar lined epithelium in response to acid reflux. Definitive eradication of this condition is difficult; thus, it is one of the few known premalignant conditions that is left in *situ*. Endoscopic surveillance is recommended in patients with Barrett’s oesophagus to map disease evolution and detect disease progression to dysplasia and oesophageal cancer, should it develop, as in early stages endoscopic management is still possible with a curative outcome.

During endoscopy, Barrett’s oesophagus is identified by the salmon-coloured mucosa compared to the more whitish appearance of the squamous epithelium. The widely established Prague classification indicates the circumferential length (C) and the maximal length (M) of the extension of Barrett’s epithelium from the top of the gastric folds into the distal oesophagus (**Figure 1**). The Prague classification is used as risk stratification tool to determine the interval for surveillance endoscopy.^3^ It is widely recommended in US, European and British guidelines as the optimal clinical classification tool for Barrett’s oesophagus;^4-7^ however only 22% of US gastroenterologists report it routinely.^7^ By nature it is minimally quantitative, subjective and subject to operator dependence, with difficulties in determining the “top of the gastric folds” due to differences in insufflation.

**Figure 1:**
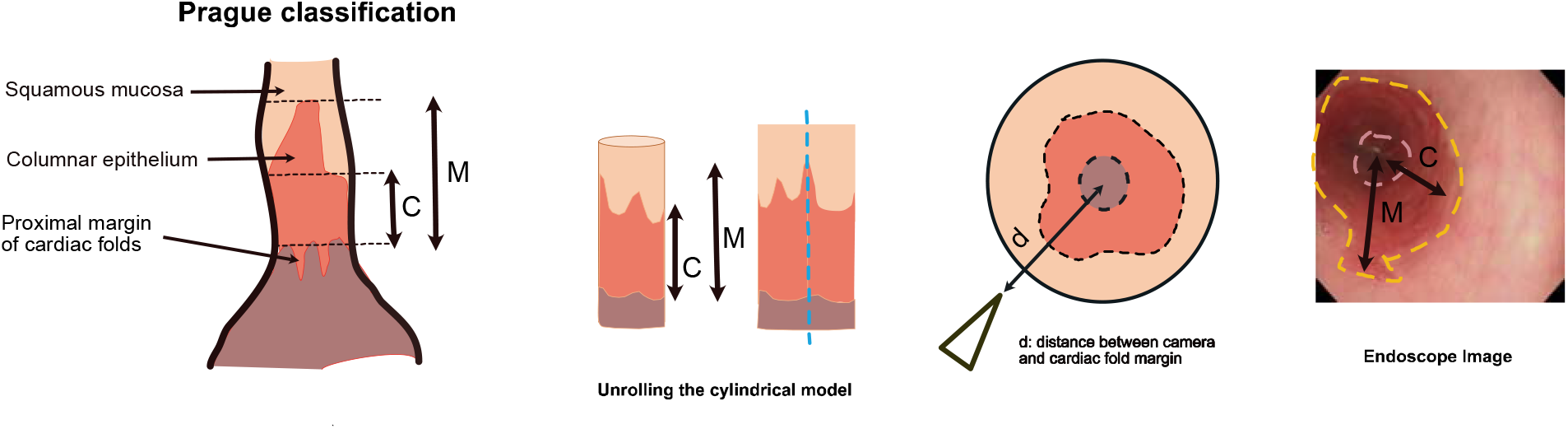
Prague classification. Oesophageal squamous and columnar cell linings depicting Prague C&M measurements taken from top of gastric folds up to the squamo-columnar junction in 3D (A) and 2D (B) are shown.

For patients with a long Barrett’s oesophagus segment ≥ 3 cm the annual progression rate to adenocarcinoma is significantly higher (0.25% per year) than for short Barrett’s length < 3 cm (0.07% per year).^8^ Therefore, the guidelines of the British Society of Gastroenterology recommend 2-3 year surveillance intervals for long Barrett’s oesophagus (>3 cm) and longer intervals (3-5 years) for short Barrett’s oesophagus with similar length based recommendations form the ESGE.^4,6^ Islands of columnar lined epithelium are ignored in the Prague classification but are encountered in a third of patients with Barrett’s oesophagus; in about half of those the islands are located proximal to the farthest extent of the Barrett’s segment and can be large especially after radiofrequency ablation.^9^ Barrett’s islands can also harbour dysplasia or EAC and their histology upgrades the overall Barrett’s epithelium dysplasia grade in 15.7% of cases.^9^

As current endoscopic surveillance programs are costly, time consuming and poorly adhered to, better risk stratification of patients with Barrett’s oesophagus to tailor surveillance recommendations is highly desirable. To date, automated, quantitative assessment of the Barrett’s length and area for risk stratification, or for monitoring the response to ablative therapy by comparing pre- and post-treatment extension is not available. Furthermore, our quantitative understanding of the temporal evolution of Barrett’s oesophagus and response to treatment is still limited. The cell-of-origin of the metaplastic epithelium is unknown although striking transcriptional similarities between cells from oesophageal submucosal glands and from Barrett’s epithelium implicate native oesophageal gland progenitor cells in a dynamic adaptive repair process.^10^ A research and clinical tool that provides quantitative assessment of the Barrett’s area and allows spatiotemporal to monitoring of topographical changes would be extremely helpful.

The aim of this study was to evaluate the possibility of assessing the Prague classification and the Barrett’s area quantification automatically by generating 3-dimensional reconstruction of the oesophageal surface from 2D endoscopic video images by leveraging camera-distances from the gastric-fold. Effectively, this 3D reconstruction provides an extended field of view which can also be used for clinical reporting and review. Building on advanced computer vision techniques, our novel automated approach is validated on simulated camera motion on a digital oesophagus model, 3D printed phantom video endoscopy data and patient data. Systematic studies presented in this paper demonstrate the accuracy and reliability of the proposed system.

## Material and Methods

### Setting and Design

This study was performed at the Translational Gastroenterology Unit at the Oxford University Hospitals NHS Foundation Trust, a tertiary referral center for endoscopic therapy of Barrett’s oesophagus neoplasia. Patients with known Barrett’s oesophagus coming for endoscopic surveillance or endoscopic treatment were included in this study. Patients undergoing upper endoscopy for dyspeptic and reflux symptoms or to investigate iron deficient anaemia served as controls. All patients included in this study provided a written informed consent for the recording of endoscopic videos and for the analysis of their clinical data. The study was approved by the local Research Ethics Committee (*REC Ref: 16/YH/0247)*.

High definition videos in white light endoscopy and narrow band imaging were prospectively recorded during endoscopy using Olympus endoscopes (GIF-H260, EVIS Lucera CV260, Olympus Medical Systems, Tokyo, Japan). Measuring and subtracting the distances from the tip of the inserted endoscope at the top of the gastric folds and at the proximal squamocolumnar margin to the incisors gives the standard circumferential and maximal length of the Barrett’s oesophagus measurements. The Prague C&M scores were reported for all endoscopies in patients with Barrett’s oesophagus. A standard biopsy forceps with known shaft diameter of 2.8 mm (Radial Jaw 4™, Boston Scientific, US) was advanced through the instrument channel into the stomach until several of the 3 mm black markers on the shaft were visible. The biopsy forceps were held in fixed position during slow withdrawal of the endoscope through the oesophagus whilst recording.

### Datasets

#### Endoscopy Patient Cohort

The endoscopy patient cohort investigated is split into three different groups:

a. Dataset 1: 68 newly diagnosed patients with known presence of Barrett’s oesophagus attending for their first endoscopy before treatment,
b. Dataset 2: 13 patients with Barrett’s oesophagus not having received endoscopic treatment between two consecutive endoscopy visits,
c. Dataset 3: 17 patients with Barrett’s oesophagus receiving endoscopic treatment for comparison of pre- and post-treatment measurements.

Detailed summary of patient data cohort is provided in **Figure 2**. The Prague score measurements were endoscopically determined by two expert upper GI endoscopists. The majority of the patients in this cohort is male (89.7%); the average age of all patients is 67.5 years. For the first visit, variable sizes of M and C scores can be observed with a mean size of 6 cm for M-value and 4 cm for C-value (**Figure 2A**, center). A majority of patients had C-value less than 3 cm length while the M-values are predominantly higher (**Figure 2A**, right). Reporting of the Prague C&M values is consistent in repeated visits with a marginal deviation only (**Figure 2B**). Pre- and post-treatment dataset provided the evidence that majority of the patients had significantly reduced Prague C&M values in the post treatment measurement (refer to C and M markers below the dashed black line in **Figure 2C**). 5 patient videos from Dataset 3 were also used for BOA measurement.

**Figure 2:**
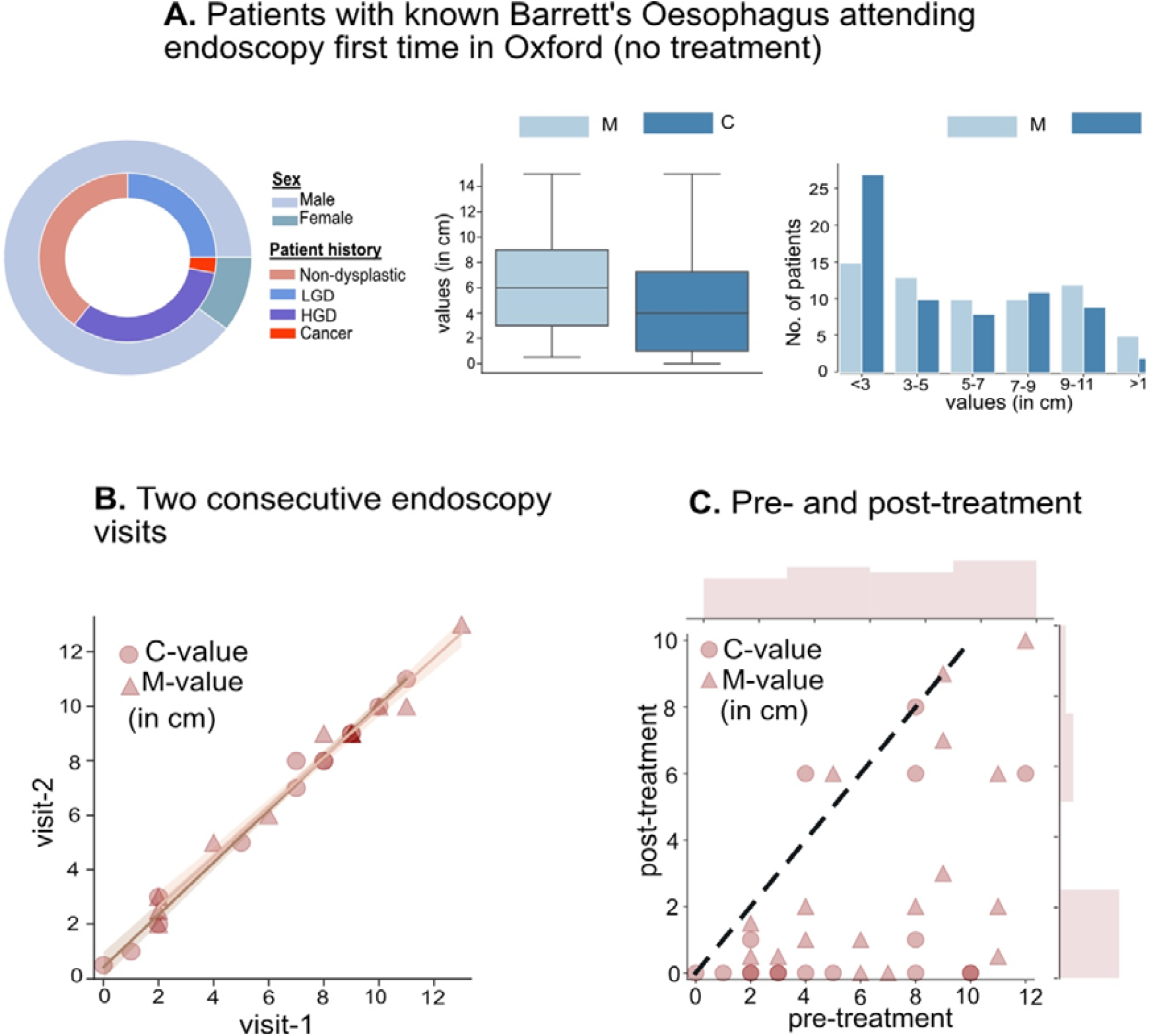
Patient cohort used for clinical evaluation study. A) 68 patients attending for their first visit (61 male and 7 female) with different histology (left, second ring of pie-plot) and subsequent treatment. Prague C&M scores presenting variation in Barrett’s length (center) and patient numbers at different C and M lengths (right). B) C and M scores measured at two consecutive visits after an average of 6 months (without treatment) is shown for 13 patients. Solid lines represent the best-fit lines between the measurements. C) Prague C&M scores of 17 patients before and after treatment. The dashed line (black) marks no changes between two measurements; most post -treatment scores show reduction in Barrett’s extension

#### Simulated Endoscopy Data

A 3D phantom model of 18.75 cm length and 2 cm internal diameter was printed using the information derived from the CT images of a reconstructed oesophagus and oesophageal endoscopy videos. Barrett’s oesophagus patterns were printed on the inside of the printed phantom using dark pink-colour silicon coating and normal squamous area was represented by light pink color (see Supplementary Figure 1 B). A third-party software, Meshmixer^11^ was used to design an unflattened oesophagus. Subsequently, the 3D model was then used to simulate endoscopy exams with known camera positions and 3D trajectories with a single light source depicted as a quadratic fall-off illumination intensity.^12^ Known camera positions and motion paths, accurate depth maps for multiple trajectory and lighting illumination scenarios were created using publicly available 3D animation software “Blender”. More than 8,000 images with corresponding ground truth depth maps for 8 different camera trajectories that include spiral, random, straight and zigzag paths were generated. A variety of different viewing angles and illumination intensity were used to mimic a real-world endoscopy system. The acquired images were 3 channel (RGB) and depth maps were 1 channel data of size 256 x 256 pixels. **Supplementary Figure 1A** shows the images acquired from different camera trajectory paths and their corresponding depth maps (distance-from-camera) in cm. To address tissue deformations and varying spatial morphology during oesophageal peristalsis, 5000 simulated colon data from well-established endoscopic depth prediction on digital 3D phantoms were also used for training purposes.^13^

To evaluate the quantification and 3D reconstruction of Barrett’s oesophagus a 3D printed phantom model of an oesophagus with salmon-coloured painting of simulated BOAs (**Supplementary Figure 1B**) was used. 10 different endoscopy trajectory videos were acquired with the same gastroscope used for patient examination. Precise ground truth measurements for Prague C&M and island lengths were acquired with Vernier callipers (**Supplementary Figure 1B**). BOAs and three painted island areas in the phantom model were measured against mm grid paper and this served as ground truth measurements.

### Artificial Intelligence for Computer-Aided Barrett’s Quantification System

The primary goal of our Barrett’s quantification system is to assist endoscopists to acquire robust and reliable Prague C&M scores automatically. The system also computes Barrett’s oesophageal area (BOA) that can be a helpful indicator for measuring risk in patients with large island segments. In addition, by mapping 2D video images to a 3D reconstruction we present a novel approach of performing a comprehensive risk analysis of Barrett’s patients which is not possible today.

**Figure 3A** illustrates the complete artificial intelligence (AI) system design of our proposed Barrett’s quantification system. The system measures the Prague C&M scores automatically leveraging the learnt camera distance estimation. A deep learning-based depth estimator network (**Supplementary Figure 2**) was trained with 10,000 simulated images (6000 oesophagus, 4000 colon) with known (simulated) distance-from-camera (depth) measurements (**Supplementary Figure 1A**). The technical challenge of accurately quantifying the Barrett’s length from a complex 3D surface as well as design of the estimators for depth and pose are described in the **supplementary material Section 1.1**.

**Figure 3:**
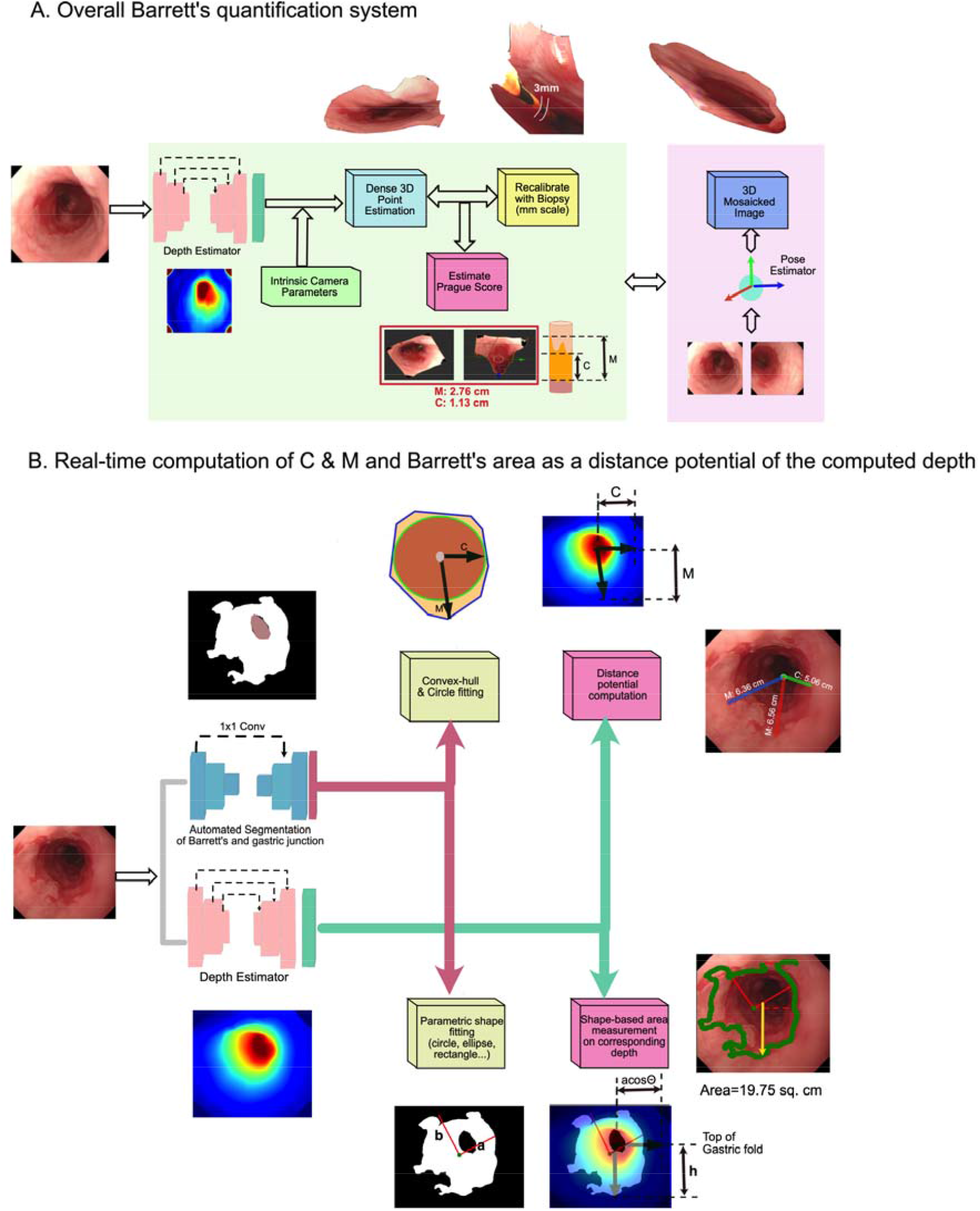
Barrett’s quantification system. 3D reconstruction, Prague C&M score measurements and extended 3D mosaicked surfaces are the output results of the proposed system. A) Overall system design for all outputs (offline). B) Real-time computation of Prague C&M criteria and BOA measurements directly from the depth estimation (online). The method involves segmentation of Barrett’s area and computer vision-based shape fittings. On the right top, C and M measurements and on the right bottom is the BOA measurement.

For 3D point projection, the intrinsic camera parameters were measured offline using checkerboard pattern images acquired by the endoscope GIF-H260 Olympus (used in this study).^14^ For large C and M values, standardised markers on biopsy forceps can be used as internal reference to recalibrate for real-world mm-scale measurements (for more details, please see **Supplementary material Section 1.1.4**).^11^ Here, we focus on real time Prague C&M estimation from depth and Barrett’s area measurements. **Figure 3B** demonstrates the analysis pipeline for our real-time Prague C&M criteria and BOA measurements. For both designs, a light-weight model is used for Barrett’s area and gastric junction segmentation (**Supplementary material 1.3**) followed by shape fittings methods in computer vision and projection of depth estimations computed from our trained depth estimator model.

For C and M measurements (**Figure 3B**, top), the centroid of the fitted circle provides the center from which the distances are measured (ideally lies in the close proximity of the gastric junction). The radius of the circle and the maximal length of the fitted convex hull projected on the predicted depth map provides the Prague C&M measures, respectively. For area measurements (**Figure 3B**, bottom), parametric shape fitting is used to compute the surface area with the corresponding depth. These computational analyses depend on the following assumptions:

1. Oesophagus is sufficiently insufflated for a fraction of second and the gastric folds are visible, and
2. Endoscopic camera is held nearly perpendicular to the gastric junction.

In the case of violation of above assumptions such as during large Barrett’s segments and invisible gastric folds, we adhere with the quantification performed by our system in **Figure 3A** which performs mosaicking of the predicted depths and images (see **Supplementary material Section 1.1.4**). In all cases, the computed depth maps allow for an efficient 3D reconstruction.

### Evaluation Criteria

#### Quantitative Evaluation Metrics

To demonstrate the accuracy of the system against acquired ground truth measurements on both digital and printed phantom, and reliability tests conducted against expert measurements we used the following metrics:

I. Computed depth predictions from our depth estimator are evaluated against ground-truth depth maps on the synthetic dataset (refer Supplementary Figure 1A) on 3000 test data using established standard metrics. Here, 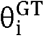 and 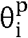 represents ground-truth and predicted depths, respectively.
  a. Relative Error (Rel.): 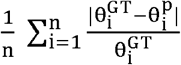
  b. Root Mean Square Error (RMSE): 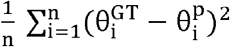
  c. Root Mean Square Log Error (RMSE Log): 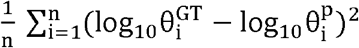
II. (We use relative error, and RMSE measures to quantify the maximal diameter of an island, C and M, and BOA measurements on the phantom endoscopy data compared to the direct measurements from the painted Barrett’s oesophagus in the phantom as ground truths.
III. For the evaluation of automated Prague C&M on patient data from video endoscopies, the absolute difference is reported between the documented Prague score by expert upper GI endoscopists and measurements obtained from the proposed quantification system.

### Statistical Analysis

In the absence of reference data from previous studies on Barrett’s quantification, no formal sample size calculation was carried out. Endoscopic Prague C&M quantification was carried out independent to the simulated data measurements. For this we grouped first visit patients according to the reported C and M lengths. Due to the small number of patients for Dataset 2 and Dataset 3, unlike for Dataset 1 we quantified each as a single individual group to increase the statistical power of the analysis. The concordance between Prague C&M scores assessed by the expert upper GI endoscopists and computer-aided quantitative measurements was assessed using Cohen’s Kappa, Kendall’s tau, and Spearman correlation. Paired t-test was used to compute the significance between our automated Prague C&M measurements compared to the reported values. For this p-values greater than 0.05 were considered statistically non-significant.

## Results

### Validation on simulated dataset

The errors in the predicted depth maps compared to the ground truth depths were quantified on 2000 simulated oesophageal images (test data) rendered from a virtual endoscopy on a digital 3D oesophagus model using third-party blender software (**Supplementary Figure 1 A**). Five different endoscopy trajectories with three different lighting settings were used to generate the test data. It can be observed in **Supplementary Table 1** that the proposed method obtained minimal errors for most trajectory data and reported the least average RMSE error of only 0.41 mm which is 1.85 mm less compared to classical FPN networks.

### Validation on phantom endoscopy dataset

The results for the automatic quantification of C & M values, maximal island diameter, and BOA from the oesophageal endoscopy videos done on the 3D printed phantom is shown in **Table 1**. The proposed method achieves more than 95 % average accuracy (4.2 % relative error) and an average deviation from the ground truth of only 1.80 mm. In addition, the RMSE error was estimated to be 2.50 mm confirming a substantial agreement (k = 0.72 and r_s_ = 0.99) with the ground-truth measurements. **Table 1** also demonstrates the validation of the Barrett’s area quantification using our proposed system (**Figure 3 B**). It can be observed that marginal difference error (least) was obtained for Barrett’s area A1 and island 1. We attribute this to the fact that the original silicon model was printed with distinct salmon colour for the Barrett’s area A1 and island1 which did not affect our segmentation and depth estimation. However, the additional Barrett’s areas (A2, A3, and islands 2-3) were painted with water colours resulting in less distinct margins; the errors are therefore higher. However, the average RMSE is only 1.59 cm^2^ with a correlation of 0.94 and kappa of 0.42 (moderate agreement).

**Table 1.** Automated Barrett’s length and area quantification using endoscopy data acquired from the 3D printed phantom: Lengths and areas of differently shaped painted Barrett’s epithelium in the phantom were measured using Vernier callipers and mm-scale grid paper. The automated measurements in the phantom from the proposed system are reported. Ma and Mb correspond to M7 and M6, respectively (see **Supplementary Figure 1B**). Similarly, the automated area measurements for three different Barrett’s and island paintings areas are shown.

| Phantom Endoscopy Dataset | Barrett's markers | Measurements |  | Errors |  | RMSE | Agreement |
| --- | --- | --- | --- | --- | --- | --- | --- |
|  |  | Ground-truth | Automated measurement | Abs. diff. | Relative error (%) |  |  |
| Length measurements (in cm) | Ma | 7.07 | 6.95 | 0.12 | 1.70 | 0.25 | Spearman rank correlation = 0.99<br>Cohen's Kappa = 0.72 |
|  | Mb | 6.90 | 6.37 | 0.53 | 7.68 |  |  |
|  | C | 2.3 | 2.12 | 0.18 | 7.80 |  |  |
|  | Island 1 | 1.99 | 2.05 | 0.06 | 3.01 |  |  |
|  | Island 2 | 1.22 | 1.21 | 0.01 | 0.82 |  |  |
|  | **Overall (average)** | **3.89** | **3.74** | **0.18** | **4.20** |  |  |
| Area measurements (in sq. cm) | Barrett's A1 | 62.50 | 62.01 | 0.49 | 0.78 | 1.59 | Spearman rank correlation = 0.94<br>Cohen's Kappa = 0.42 |
|  | Barrett's A2 | 68.50 | 65.03 | 3.47 | 5.06 |  |  |
|  | Barrett's A3 | 76.50 | 78.07 | 1.57 | 2.05 |  |  |
|  | Island 1 | 3.50 | 3.61 | 0.11 | 3.14 |  |  |
|  | Island 2 | 3.70 | 3.10 | 0.60 | 16.21 |  |  |
|  | Island 3 | 3.00 | 2.55 | 0.45 | 15.00 |  |  |
|  | **Overall (average)** | **36.28** | **35.72** | **1.11** | **7.04** |  |  |

### Validation on patient dataset

**Figure 4** shows the comparison between C and M scores determined by the upper GI expert endoscopists and the automated measurements from our proposed system in our patient cohort. Even though large deviations are observed for the patient group with short segment Barrett’s oesophagus of less than 3 cm in C and M values (p-value < 0.05; **Figure 4 A**); for almost all other groups the p-value is non-significant. The box-plots for all patients in this cohort show similar median, minimum and maximum deviations. **Table 2** illustrates the results from the proposed AI system and from the expert endoscopists in determining the Prague classification in the cohort of patients with Barrett’s oesophagus. The mean deviation (absolute difference) in measurement for both C and M for each group is less than 7 mm with an overall deviation of 4.5 mm and 6.0 mm respectively for Prague C&M values. The overall agreement between the C and M values reported by the expert upper GI endoscopists and the automated AI system is expressed with k of 0.59 and 0.50 for C and M, respectively, and over 90% τ and r_s_ on this dataset.

**Table 2.**
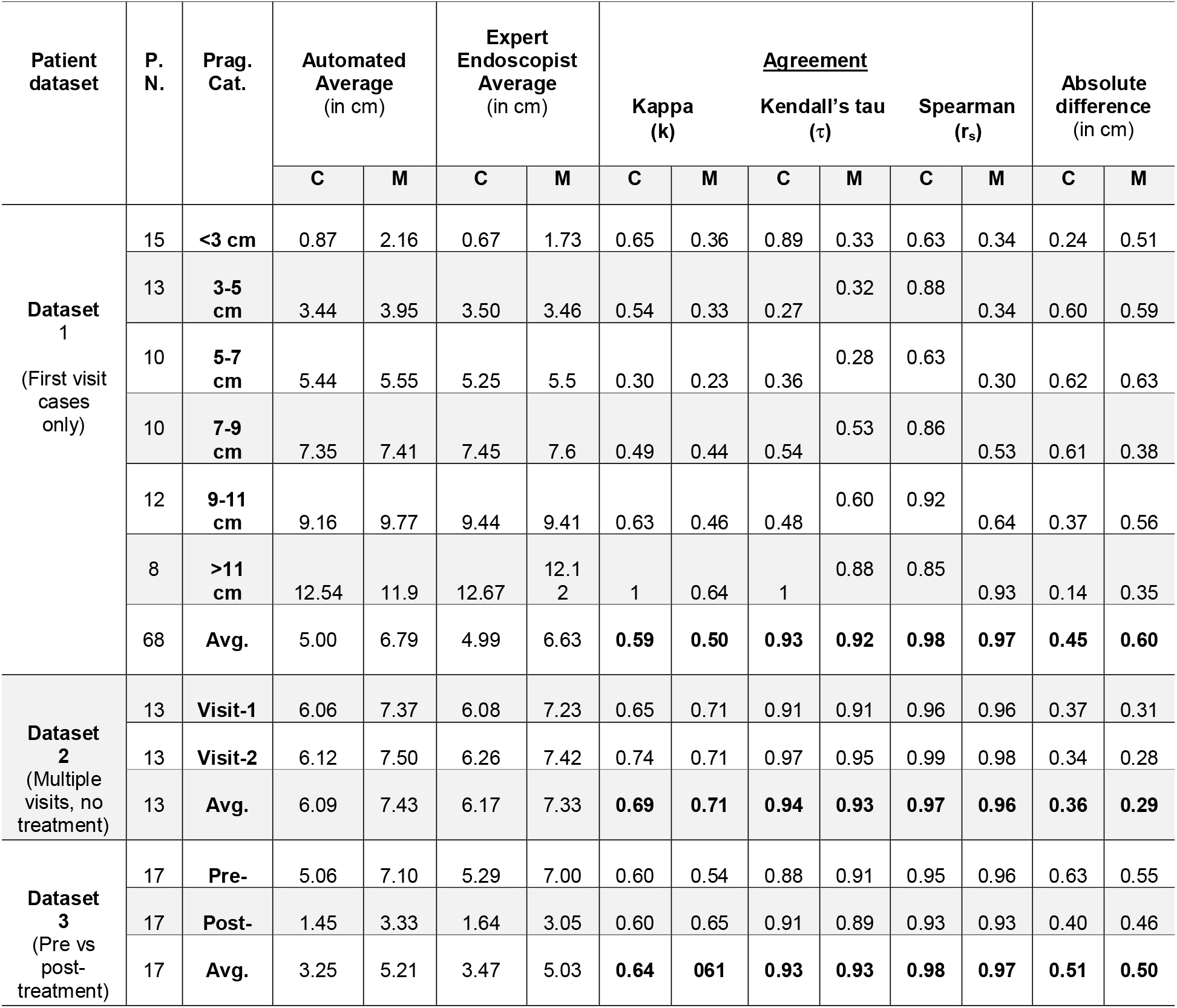
Automated quantification of. Prague C&M scores from real patient video data compared with the endoscopists’ measurements. Statistic measures to analyse the concordance between the automated and the measurement reported by the endoscopist are also provided. P.N = total patient number; Prag. Cat. = Prague Category; Avg. = overall Average.

**Figure 4.**
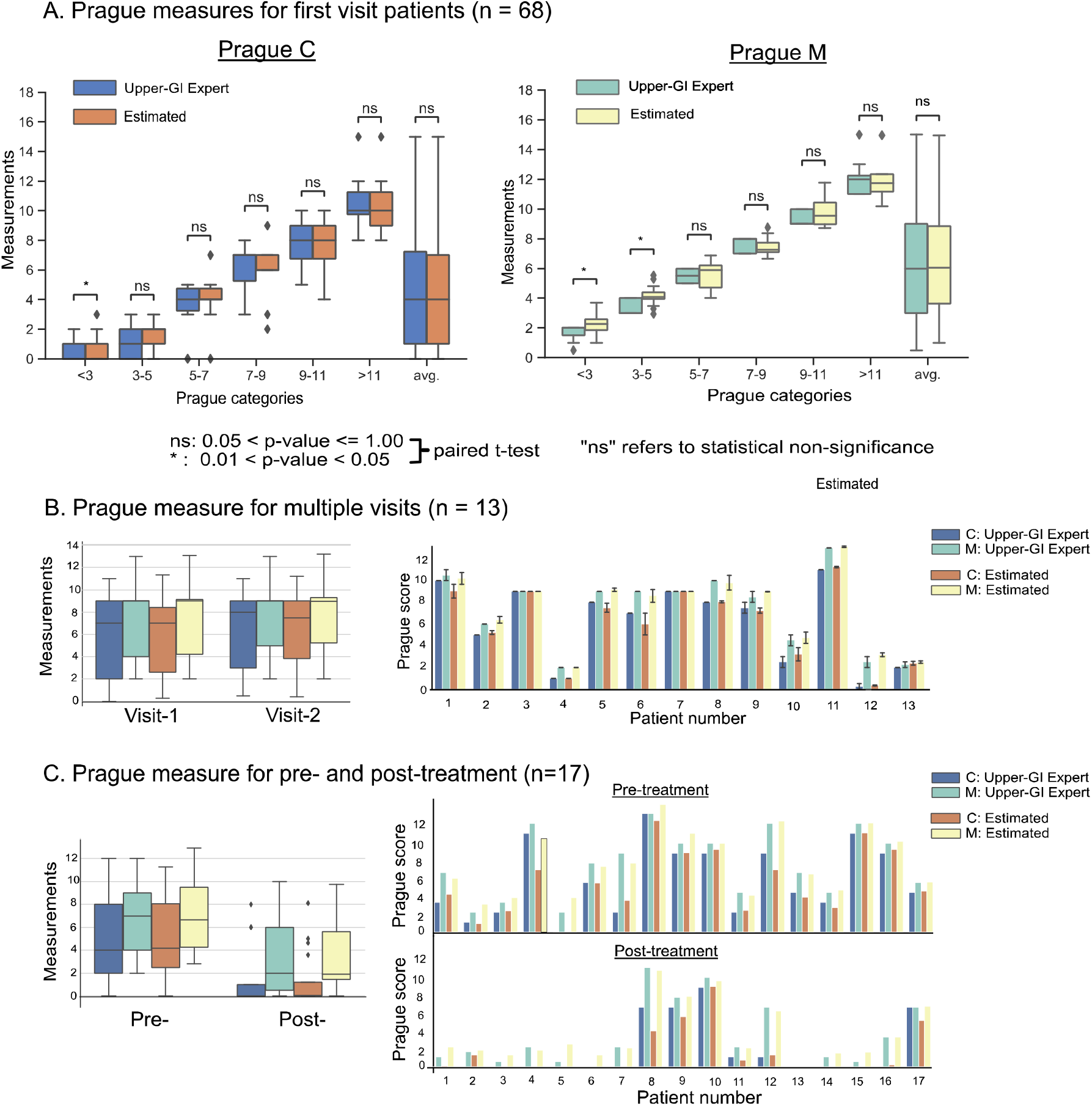
Measurement of Barrett’s C and M scores in 98 patients: Comparison between Prague C&M scores reported by expert upper GI endoscopists and the automated measurements from our proposed system. A) From left to right, Prague C&M score comparison for various length categories (less than 3 to over 11 cm). Non-significant (p > 0.05) is represented as ns. B) Expert upper GI endoscopists and automated Prague scores measurements for two consecutive patient visits. It can be observed that no significant changes were observed (also refer to error bars for each patient on the right), and our system correlated well with experts. C) Pre-treatment and post-treatment measurements, also shown for individual patient on the right. On left, the box plot shows decreased measurements for both C and M measures computed by the expert and our proposed automated system.

Error bars in bar plots demonstrating C and M values for dataset 2 (13 patients attending for repeated visits without receiving treatment) in **Figure 4 B** show only minor changes in measurements which is evident from the **Table 2** that shows 3.6 mm and 2.9 mm mean deviations for C and M values. Kappa agreement (k) > 0.7 (substantial) and > 90% for rank correlation measures (τ and r_s_) can be observed.

**Supplementary Table 2** demonstrates the applicability of the area measurement to quantify the efficacy of the ablation therapy in 5 patients with Barrett’s oesophagus. It can be observed that even though the C and M measures are reduced for all 5 patients, however, in most cases the residual Barrett’s areas/islands are more than 10 cm^2^ after the first ablation and in one case this is as large as > 26 cm^2^ although both C and M values are zero.

### Qualitative assessment of the system outputs

**Figure 5 A** shows the predicted depths on simulated test dataset for oesophageal surfaces. It can be observed that the predicted depth maps are smooth and continuous, where the camera pose is one of the dependent variables. The absolute difference between ground truth and predicted maps represents the error in our distance prediction shown on the fourth column. All errors are less than 5.0 mm for the oesophageal model. The predicted depth maps and the corresponding 3D views with their respective C and M values for patient endoscopy frames are shown in **Figure 5 B**. The Barrett’s area segmentation, convex hull fitting and mapping of predicted depths as illustrated in Figure 3B has been applied for near real-time visualisation of the Prague C&M estimates for our high-resolution video endoscopy (3.5 milli-second, nearly 27 frames-per-second). Similarly, **Supplementary Figure 3** represents the automated measurement of Barrett’s area pre- and post-treatment in two different patients. It can be observed that the large island post-treatment in the first patient recorded an area of 26.73 cm^2^. It also illustrates that the choices of shape fittings is based on the shape of the segmented Barrett’s surface.

**Figure 5.**
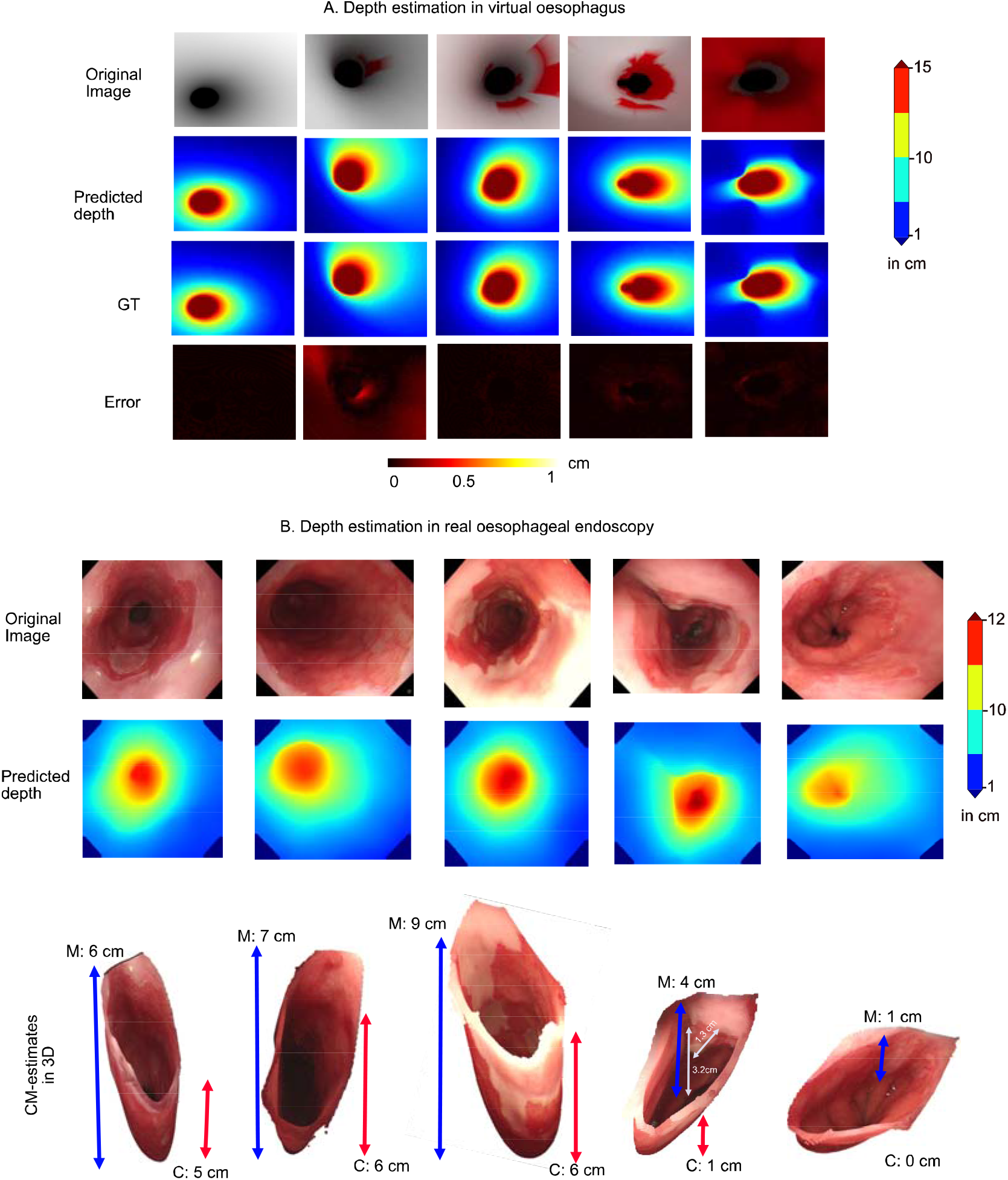
Depth-from-camera estimation and Prague C&M measures. Comparison of estimated depth with respect to the predicted depth for virtual oesophagus model (A). Error in prediction is presented as the difference between the predicted and ground-truth (GT) distances. Results for the depth estimation on real gastroesophageal endoscopy is presented in (B) showing a range of 0-12 cm distance measured from camera. The Prague C&M values in 3D reconstructed oesophagus is also shown (scaled for visualisation). 3D oesophagus can be interactively rotated, translated allowing measurements of size and distance also from encountered lesions (e.g. the small nodule in the image in the 4^th^ column is 1.3 cm in size an approximately 3.2 cm from the gastric fold) or biopsy spots in relation to the top of the gastric folds and the squamocolumnar margins.

## Discussion

The proposed methodology holds the promise of providing a quantitative, reliable, and automated assessment of Barrett’s length and area suitable for routine clinical use. Although well-established, Prague classification is only a rough estimate for the extent of the Barrett’s epithelium. Excluding Barrett’s islands, the Prague classification provides only a surrogate measure and not a true quantitative analysis of the total area of Barrett’s epithelium. In this study, we developed and tested a real-time AI system that automatically identifies, delineates and quantifies Barrett’s epithelium by recognising the typical landmarks at the top of the gastric folds and the proximal squamocolumnar margin.

We present a systematic analysis of the measurement output by comparing it against reference data acquired by imaging a purpose built phantom as well as comparing it with estimates provided by expert endoscopists. Reliability of the proposed depth measurement algorithm against various camera trajectories and lighting conditions, and the least average RMSE error less than 0.5 mm in the simulated virtual endoscopy data (see Supplementary Table 1 and Supplementary Figure 1 A) suggests precise measurement capability of the designed AI model. The Prague C&M score reported by the upper GI specialists correlated well with the automatically measured values on patient data with an average standard deviation of ± 4.5 mm and ± 6.0 mm for C and M values, respectively, showing moderate (k = 0.50) to substantial (k = 0.71) agreement with expert upper GI endoscopists for both short and long Barrett’s segments patients (**Table 2**). Exceeding this, the quantitative validation on the endoscopic phantom video data demonstrated over 95 % accuracy with only ± 1.8 mm average deviation, k = 0.72 and r_s_ = 0.99 from the available precise ground-truth measurements (**Table 1**); this implies that the computer aided measurement of the Prague scores is more precise than the measurement by the expert upper GI endoscopists during endoscopy. **Figure 4** B shows repeated measurements of Prague C&M for 13 patients during two visits where no change in Barrett’s length were observed by the system consistent with the expert finding. Similarly, for patients undergoing one session of radiofrequency ablation (**Figure 4 C)**, a reduction in the length of the Barrett’s segments (mostly C) at the next visit were marked by the system. Both of these reproducibility tests provide evidence of internal validity and responsiveness.

Existing clinical studies indicate that an accurate and more systematic assessment of the Barrett’s area will be of clinical value. Anaparthy et al.^15^ demonstrated that with every centimetre increase in M-score of Barrett’s, the risk of progression to high-grade dysplasia or EAC increases by 28 % (p = 0.01). Barrett’s segment >= 3cm showed significantly greater prevalence of dysplasia (23% vs 9%, p = 0.0001).^16^ A recent meta-analysis demonstrated that non-dysplastic short segment Barrett’s oesophagus has significantly lower rates of neoplastic progression than long segments.^17,18^ It is thus critical to report precise measurements. In addition, measurement of the BOA should be incorporated in reporting to measure risks more reliably. The built technology allows for mm-scale measurements of both Barrett’s lengths and areas. Sharma et al.^19^ demonstrated that recurrence of intestinal metaplasia after ablation therapy in the form of islands is common showing 8%-10% per patient-year by some studies.^20-21^ To address this issue, 3D reconstruction of Barrett’s can be a step forward for effective follow-up of the mucosa.

By applying our system to the quantitative analysis of a patient’s response to therapy we demonstrate that measuring of the entire Barrett’s provides further evidence of tangible improvements over the commonly used Prague scores. A patient who received radiofrequency ablation with reported Prague C0M0 score had considerably large untreated BOA (see **Supplementary Figure 3**). Such insular areas could potentially harbour cancer or dysplasia.^9^ It is therefore eminently important to build technologies that can provide Barrett’s area for quantification of therapy response. The validation and reliability tests on three phantom endoscopy video data (with known measurements) showed the efficacy of our proposed BOA measurement for which only 1.11 cm^2^ average deviation was observed compared to the ground-truth BOA with moderate kappa agreement (k = 0.42) and strong Spearman rank correlation (r_s_ = 0.94). The study included three different island sizes and three differently shaped Barrett’s areas.

In addition to enabling precise and systematic measurements of the Barrett’s area, the proposed 3D surface reconstruction is likely to revolutionise our way of reporting Barrett’s surveillance endoscopy and corresponding histology requests. By documenting biopsy locations and encountered pathology (see **Figure 5 b, 4**^**th**^ **column**), it provides a natural linkage between endoscopy and any further histopathological assessment. This compact representation can also be the basis for an additional specialist review. The quantification of the entire area of Barrett’s epithelium is plausibly a better tool for risk stratification to measure progression to Barrett’s neoplasia than the currently used extension in length. Barrett’s area in combination with dysplasia grade or quantified pit pattern morphology might guide determination of intervals for endoscopic Barrett’s surveillance in future. Up to now, there is no research tool available to investigate and quantify the emergence of Barrett’s oesophagus over time.

This novel AI system will allow to monitor temporal morphological changes of Barrett’s oesophagus during development or possible regression and in response to any treatment. Quantification of the Barrett’s area can be used to assess treatment efficacy after ablative treatment of dysplastic Barrett’s oesophagus such as radiofrequency ablation, cryoablation, argon plasma coagulation or stepwise endoscopic resection.

Limitations of this study, this is a single center pilot study which needs further evaluation in the clinical, non-expert endoscopy setting. We have not evaluated the automatic quantification of Barrett’s extension and reconstruction from endoscopy against measurement after surgical oesophagectomy. However, the surgical resection specimen will also be subjected to shrinking artifacts and contractions and not present the true in-vivo dimensions.

In conclusion, we present a deep learning-based AI system that reliably quantifies the extension of Barrett’s epithelium in real-time endoscopy. Rather than requiring any extensive hardware mortifications, this technology is very accessible and only requires a minimal amount of training. It holds the potential of enhancing endoscopic screening programmes by providing quantitative and objective data that can be used for review and the assessment of disease progression.

## Data Availability

The authors confirm that the data supporting the findings of this study are available within the article and its supplementary materials.

---

**Supplementary Table 1.**
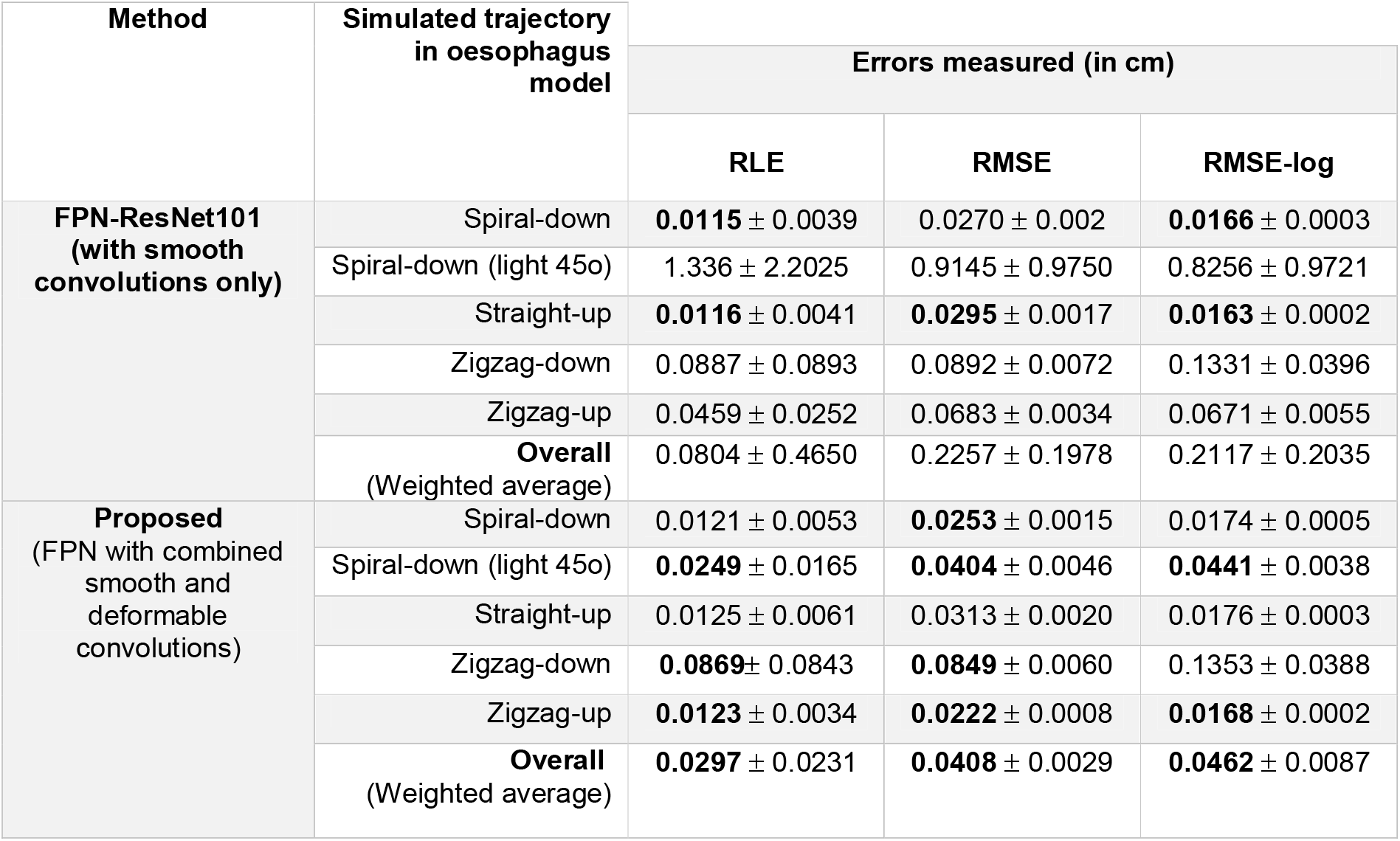
Error measured for the predicted depth maps on virtual oesophageal endoscopy data: Errors are presented in cm. Best results are highlighted in bold.

**Supplementary Table 2.** Automated Barrett’s area quantification for pre- and post-treatment for 5 patients. No ground-truth measurements are available. Evidence for large Barrett’s area (>10 sq. cm) post-treatment are underlined (also see **Supplementary Figure 3**).

| Patient ID. | Pre-treatment |  |  | Post-treatment |  |  |
| --- | --- | --- | --- | --- | --- | --- |
|  | C-value (in cm) | M-value (in cm) | Area (in cm<sup>2</sup>) | C-value (in cm) | M-value (in cm) | Area (in cm<sup>2</sup>) |
| 1332 | 3 | 6 | 65.00 | 0 | 1 | 9.00 |
| 2006 | 2 | 3 | 47.92 | 0 | 0.5 | 5.21 |
| 2021 | 10 | 11 | 83.01 | 0 | <u>2</u> | <u>11.87</u> |
| 3080 | 4 | 6 | 62.05 | <u>0</u> | <u>0</u> | <u>26.73</u> |
| 3164 | 3 | 4 | 34.62 | 0 | 1 | 9.01 |

**Supplementary Figure 1:**
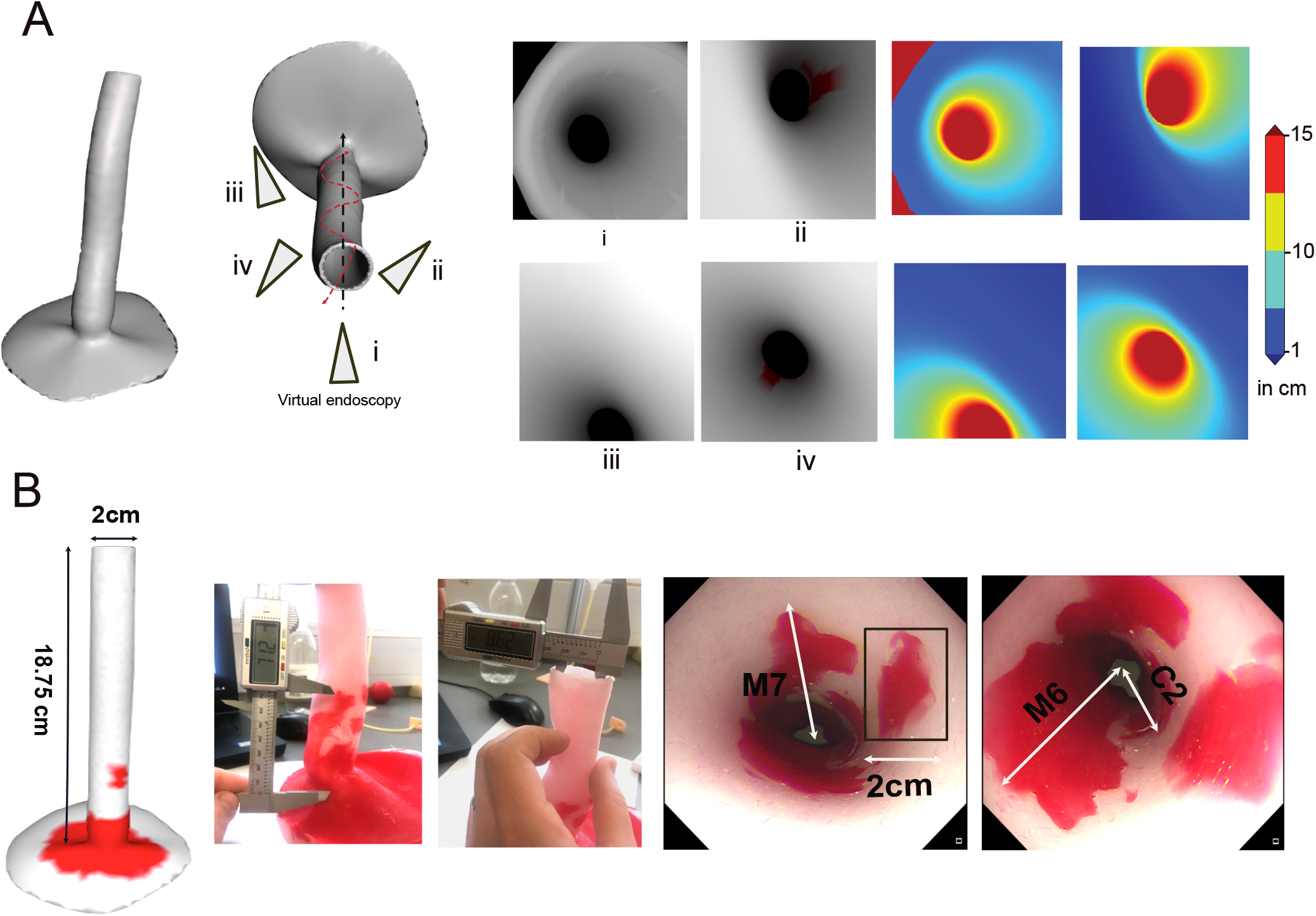
(A) **Simulated depth maps in a virtual 3D oesophagus model**. On left: Camera trajectories (i-iv) representing straight and spiral camera motion. On right: Endoscopic images and their corresponding depth map estimation for each trajectory shown on left (distance from endoscopy camera). (B) **Validation data acquisition:** 3D printed oesophagus phantom model with known measures for C, M (white arrows) and island (black rectangle). Endoscopy video frames are shown on the right.

**Supplementary Figure 2.**
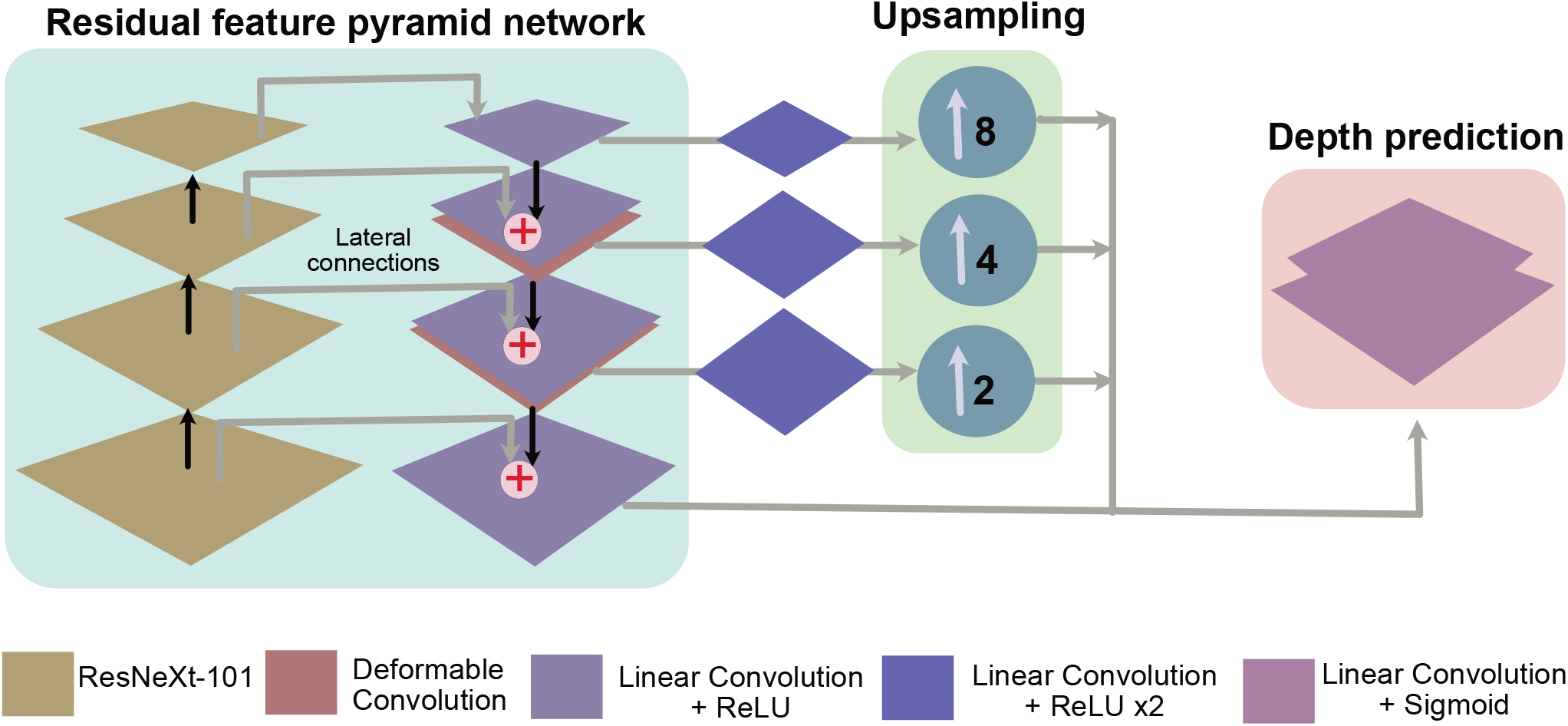
Depth estimator: Proposed deep-learning framework for estimating camera-distances from the target region (depth estimation).

**Supplementary Figure 3.**
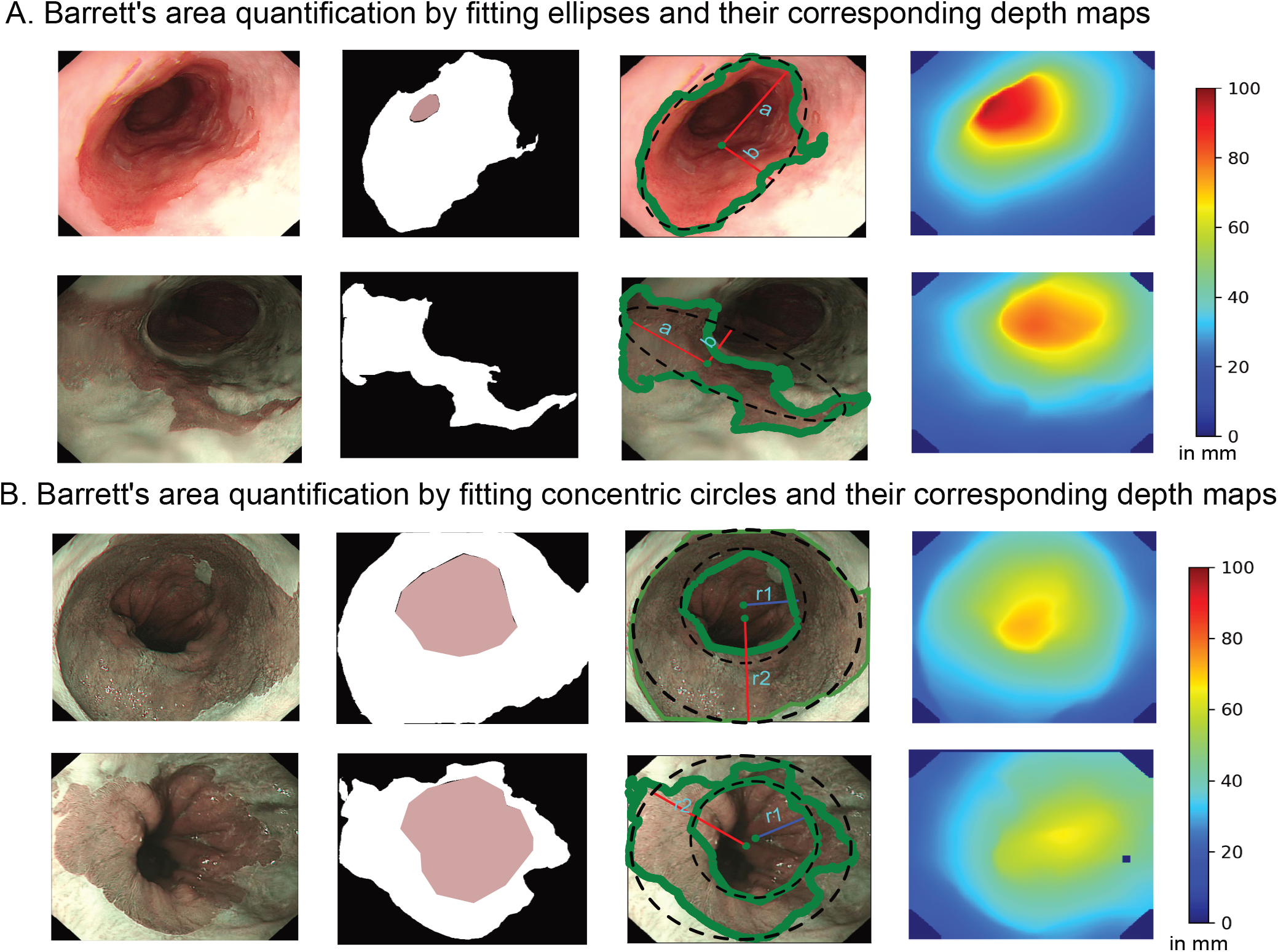
Automated measurements for Barrett’s area using different parametric shape fittings. (A) Elliptical area fitting on the segmented mask of patient Id 3080 (see Table 3, original manuscript). Top: pre-treatment area of 62.05 sq. cm and bottom: post-treatment area of 26.73 sq. cm. (B) Circle fitting on the segmented mask of patient Id 2006. Two concentric circular area measurements are done to eliminate area around the gastric fold. Top: pre-treatment area of 47.92 sq. cm and bottom: post-treatment area of 5.21 sq. cm.

