## Supplementary material for "Artificial intelligence-driven real-time 3D surface quantification of Barrett’s oesophagus for risk stratification and therapeutic response monitoring"

**Short title:** Automated quantification of Barrett's epithelium

Sharib Ali[1,3], Adam Bailey[2,3], James E. East[2,3], Simon J. Leedham[3,4], Maryam Haghighat[1], TGU Investigators, Xin Lu[5], Jens Rittscher[1,3,6], Barbara Braden[2,3,6]

[1]Institute of Biomedical Engineering, Big Data Institute, Department of Engineering Science, University of Oxford, Oxford, UK; [2]Translational Gastroenterology Unit, Nuffield Department of Medicine, Experimental Medicine Div., John Radcliffe Hospital, University of Oxford, Oxford, UK; [3]Oxford NIHR Biomedical Research Centre, Oxford, UK; [4]Intestinal Stem Cell Biology Lab, Wellcome Trust Centre Human Genetics, University of Oxford, Oxford, UK; [5]Ludwig Institute for Cancer Research, University of Oxford, Oxford, UK; [6]shared senior authorship

### TGU Investigators

Philip Allan; Tim Ambrose; Carolina Arancibia-Cárcamo; Ellie Barnes; Elizabeth Bird-Lieberman; Jan Bornschein; Oliver Brain; Jane Collier; Emma Culver; Alessandra Geremia; Bruce George; Lucy Howarth; Kelsey Jones; Paul Klenerman; Rebecca Palmer; Fiona Powrie; Astor Rodrigues; Jack Satsangi; Alison Simmons; Simon Travis; Holm Uhlig; Alissa Walsh

### Supplementary Material

#### 1.1. Geometry problem, depth and pose estimator

1.1.1. **Geometry problem:** Given estimated depth $\theta_t$ and $\theta_{t-1}$ for frames $X_t$ and $X_{t-1}$ with the position and orientation of the camera represented by camera pose $E_{t\rightarrow t-1}$, the projected 3D points[1] for a single image can be written as (1) with (x, y) as image pixel coordinates.

$$P_t^{xy} = \theta_t^{xy}.K^{-1}[x,y,1]^T, \quad (1)$$

Here, $K$ is the camera intrinsic matrix obtained by the offline camera calibration.[2] The mapping of image co-ordinates at time $t$ to $t-1$ allows to transform frame $X_t$ to $X_{t-1}$ using Eq. (1) and camera pose matrix $E_{t\rightarrow t-1}$:

$$\hat{X}_{t\rightarrow t-1}^{xy} = X_t^{\hat{x}\hat{y}}, \qquad [\hat{x},\hat{y},1]^T = KE_{t\rightarrow t-1}(\theta_{t-1}^{xy}.K^{-1}[x,y,1]^T) \quad (2)$$

Similarly, this process is repeated for $n$ frames with sufficient overlap giving an extended field-of-view for Barrett's quantification required for longer Barrett segments. At sufficient insufflation of the oesophagus, the measured depths can be leveraged to calculate the Barrett's segment lengths without any internal references.

1.1.2. **Depth estimator** network is composed of residual feature pyramid network (RFPN)[3] with ResNeXt-101 backbone pretrained on imageNet. The FPN allows to extract features and semantics at multiple scales. In order to increase the receptive field and to tackle occlusions due to local deformations we have incorporated a non-linear deformable convolution kernels[4] in our proposed model in the two middle lateral connections of RFPN together with a linear convolution. All upscaled layers on the right side of the RFPN are subsequently convolved with a sequence of linear convolution kernels and ReLU activation functions. The concatenated feature maps obtained after upsampling block is finally used to predict the depth $\theta$ using a linear $3 \times 3$ convolution filter and a non-linear sigmoid function (see shown in **Supplementary Figure 2**).[5] The used loss functions are described below in **Supplementary Section 1.2**.

1.1.3. **Pose estimator** implementation of open3D[6] was to estimate SE3 transformation (camera rotation and translation) between two image frames referred as camera pose matrix $E$ in this paper.

1.1.4. **System design**: A real-time depth estimator for Prague C & M classification and Barret's area measurement system is designed and can be directly used during the endoscopic screening or therapy procedures (see **Figure 3** of the main manuscript). The Barrett's length upto 10 cm can be measured without any difficult provided the oesophagus is sufficiently insufflated.

In cases where Prague C&M are greater than 10 cm (often rare), the system takes into account the pose estimator to reconstruct the Barrett's surface from few (usually 2-4) frames (see **Figure 3A**). While in most cases ego-motion alone serves the purpose of 3D mosaicking and Barrett's quantification. Sometime due to unclear visibility of squamo-columnar junction or the gastric folds together, standardised biopsy forceps can be used as an internal reference of the 3D reconstructed surface. This can bring a slight latency (usually 5-10 seconds) in the 3D visualisation and estimation. However, recalibration is required only once but needs to be repeated for a different camera calibration setting.

Since the system allows to print a .ply file that composite point clouds, third party softwares such as Open3D can be used to visualise the reconstructed surface in 3D during follow-ups. These softwares do not require additional hardware and can be effectively used in the same system.

### 1.2. Loss functions:

Loss function used to minimise the difference between the estimated depth $\theta_i^p$ and ground truth depth $\theta_i^{GT}$ utilises four different losses defined below:

a) *Depth loss* is computed as the root mean square error (RMSE) in log scale:

$$L_d = \frac{1}{n} \sum_{i=1}^{n} \sqrt{ln^2 \theta_i^{GT} - ln^2 \theta_i^p} \tag{3}$$

b) *Gradient loss* is the $l_1$ norm of the difference between gradient of the depth maps to penalise errors around edges.

$$L_g = \frac{1}{n} \sum_{i=1}^{n} ||\nabla \theta_i^{GT} - \nabla\, \theta_i^p||_1 \tag{4}$$

c) *Surface normal loss:* The normal vector loss computed from ground truth depth $\mathrm{n}_i^{GT}$ and predicted depth $\mathrm{n}_i^p$.[7]

$$L_n = \frac{1}{n} \sum_{i=1}^{n} \left( 1 - \frac{< \mathrm{n}_i^{GT}, \mathrm{n}_i^p >}{\sqrt{< \mathrm{n}_i^{GT}, \mathrm{n}_i^{GT} >} \sqrt{< \mathrm{n}_i^p, \mathrm{n}_i^p >}} \right) \tag{5}$$

d) *Reconstruction loss:* It is computed as the l2-norm between the reconstructed 2D images from the estimated depth maps (see Eq. (2)). This loss is computed only if two frames are fed to the network.

$$L_r = \frac{1}{n}\sum_{i=1}^{n} ||X_{t-i\rightarrow(t-i-1)} - X_{(t-i-1)}||_2 \quad (6)$$

#### 1.3. Barrett's area segmentation:

We used an encoder-decoder framework with ResNet-50 backbone and atrous separable convolutions (referred as DeepLabv3+)[8] for segmentation of Barrett's area. The hollow area inside the segmented Barrett's determined the gastric fold junction. Also, we used simple area elimination to eliminate small island like objects as post-processing step (refer to the red block of automated segmentation in **Figure 3B** of the original manuscript). The entire network was trained for 50 epochs with 327 images and validated on 47 images. All images were resized to 256 pixels x 256 pixels. Stochastic gradient descent with learning rate of 0.01 and momentum of 0.9 were used. The inference time reported was > 35 frames-per-second. The network achieved an intersection-over-union score or over 78%.
